# A high efficient hospital emergency responsive mode is the key of successful treatment of 100 COVID-19 patients in Zhuhai

**DOI:** 10.1101/2020.03.15.20034629

**Authors:** Jin Huang, Zhonghe Li, Xiujuan Qu, Xiaobin Zheng, Changli Tu, Meizhu Chen, Cuiyan Tan, Jing Liu, Hong Shan

**Author notes:** These authors contributed equally to this work (Jin Huang, Zhonghe Li). Corresponding author: Hong Shan, Jing Liu, Address for correspondence, Fifth Affiliated Hospital of Sun Yat-sen University, 52 East Meihua Rd., Zhuhai City 519000, China., (Hong Shan); (Jing Liu).

## Abstract

**Background:** Since December 2019, Coronavirus Disease 2019 (COVID-19) emerged in Wuhan city and rapidly spread throughout China. The mortality of novel coronavirus pneumonia (NCP) in severe and critical cases is very high. Facing this kind of public health emergency, high efficient administrative emergency responsive mode in designated hospital is needed.

**Method:** As an affiliated hospital of Sun Yat-sen University, our hospital is the only designated one for diagnosis and treatment of COVID-19 in Zhuhai, a medium-sized city. Novel coronavirus pneumonia department, which is administrative led by the president of hospital directly, has been established at early stage of epidemic crisis in my hospital. In NCP department, there are core members of Pulmonary and Critical Care Medicine (PCCM) specialist and multidisciplinary team. Don’t stick to national guidelines of NCP, based on professional opinion by respiratory professor and expert group, we focused on individualized treatment and timely adjustment of treatment and management strategies in working about COVID-19 patients.

**Results:** 1. High working efficiency: By Mar 02, 2020, we have completed 2974 citywide consultations and treatment of 366 inpatients, including 101 patients diagnosed with COVID-19. 2. Excellent therapeutic effect: Among 101 hospitalized patients with confirmed COVID-19, all were cured and discharged, except for one death. No secondary hospital infection, no pipeline infection and no pressure sore were found in all patients. 3. Finding and confirming person-to-person transmission characteristic of COVID-19 prior to official release conference: Strengthened protection is key point to zero infection in healthcare group and medical faculty and lower rate of second generation infectious patients. 4. Timely adjustment management and treatment strategy prior to guideline update: The first evidence of digestive tract involvement in COVID-19 has been found, and the earliest clinical trial of chloroquine in the treatment of COVID-19 has been carried out in our hospital.

**Conclusions:** In our hospital, establishment of NCP department, which is administratively led by the president of hospital directly and specialized conduct by respiratory professor, is the key to success in management and treatment of COVID-19 patients. This hospital emergency responsive mode could provide reference for other hospitals and cities in epidemic situation.

## Introduction

Since December 2019, COVID-19 emerged in Wuhan city and rapidly spread throughout China and worldwide. Up to March 3, 2020, there are 80 174 confirmed patients and 2915 deaths in 31 provinces in China. Outside of China, there are 8774 confirmed patients and 128 deaths in 64 countries^1^. In different regions and different hospitals, there are different management models for respiratory infectious diseases, so that the treatment effects are different, too^2^. At the beginning of this new epidemic crisis, the missing or insufficient hospital emergency responsive plan caused rapid spread of COVID-19 and high mortality in severe COVID-19 patients. Which further results in huge expense to fight the epidemic nationwide subsequently^3^. Therefore, some effective hospital management models need to be shared, summarized and referenced in order to stop the epidemic as soon as possible and reduce mortality.

## Method

There are 72 public medical and health institutions, including 4 tertiary hospitals in Zhuhai, a medium-sized city in Guangdong province. As an affiliated hospital of Sun Yat-sen University, my hospital is the only designated one for diagnosis and treatment of COVID-19 patients in Zhuhai. It’s responsible for the consultation and centralized treatment of patients with COVID-19. Since the first patient was admitted on January 17, my hospital started the emergency plan for infectious diseases. The emergency responsive mode was still traditional mode: patients were admitted in infectious department, with help of consultation from PCCM department and instruction from administrative department in hospital (Fig. 1). But poor treatment effect and low work efficiency had been assessed after four days, a new decision from president of hospital was made quickly.

**Figure 1.**
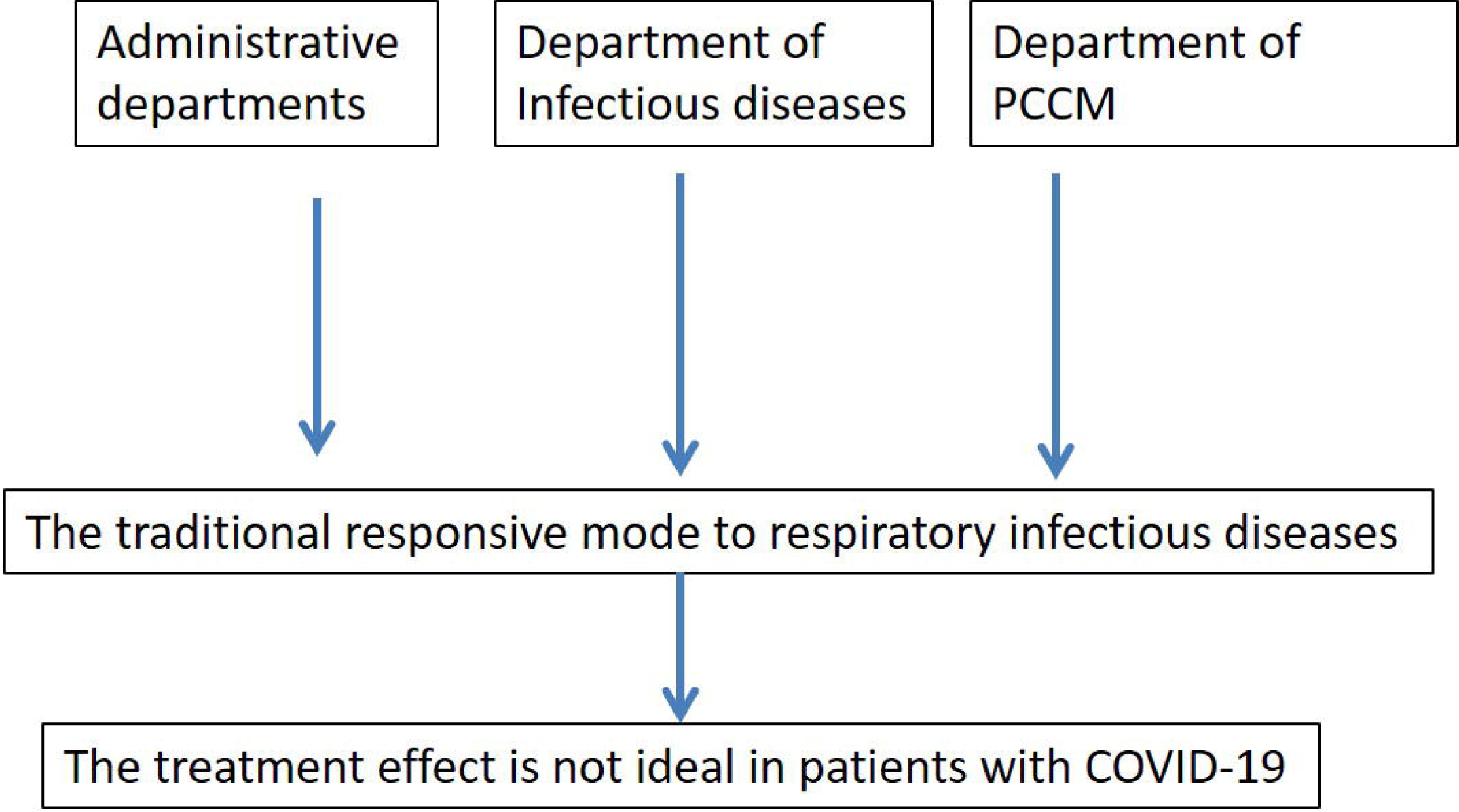
Traditional responsive mode to respiratory infectious diseases PCCM: Pulmonary and Critical Care Medicine; COVID-19: Coronavirus Disease 2019.

NCP department was established with direct administrative leading and management by president of hospital, and professional guidance by professor and faculty from PCCM. The department of infectious diseases was transferred to the periphery in charge of infection control, multidisciplinary consultation was referred for severe and critical patients, final treatment strategy was decided by professor of PCCM, and the whole hospital was mobilized to serve mainly NCP department and fight to COVID-19 crisis (Fig. 2). Don’t stick to national guidelines of NCP, based on professional opinion by respiratory professor and expert groups we focused on individualized treatment and timely adjustment of treatment and management strategies in working about COVID-19 patients.

**Figure 2.**
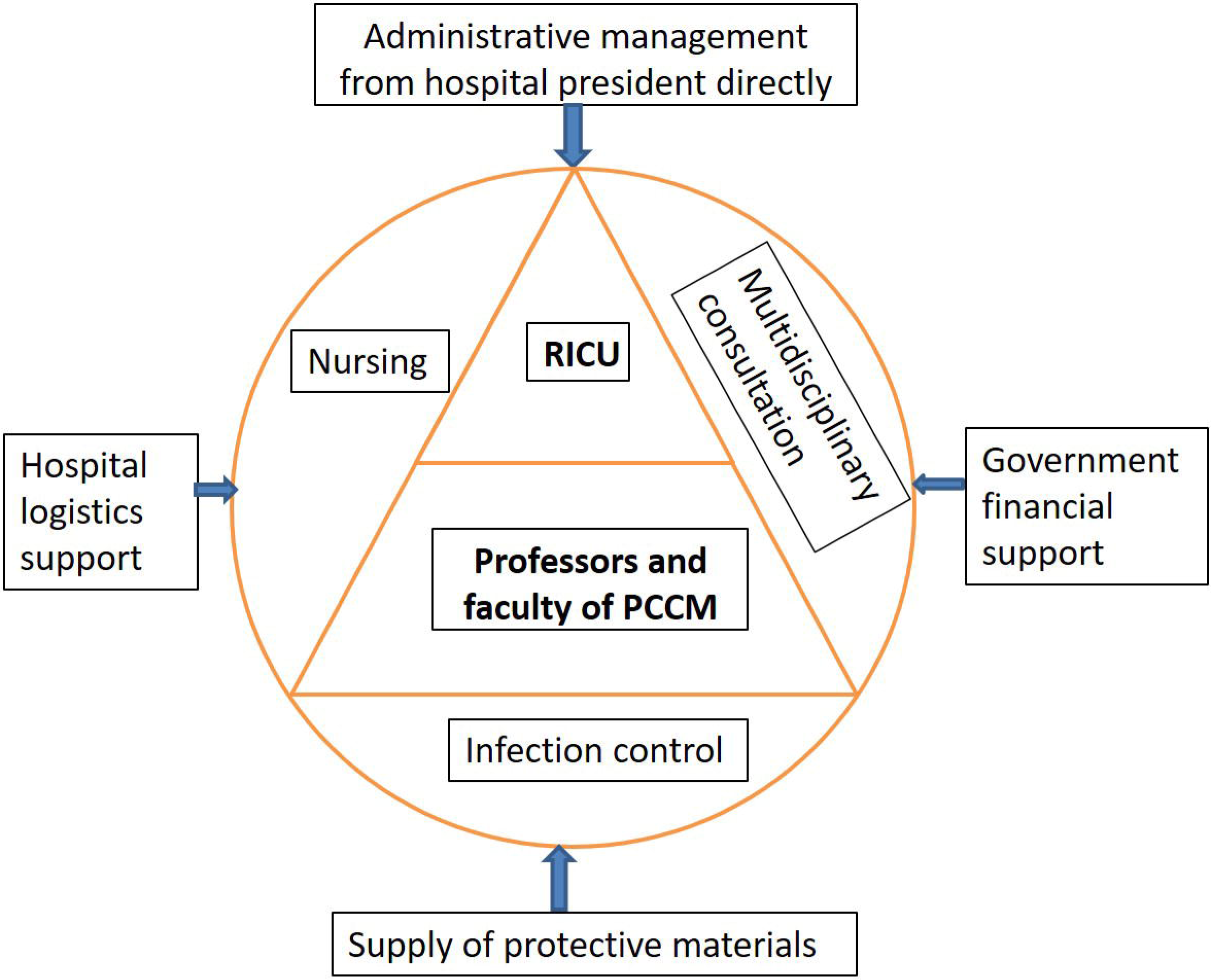
A high efficient hospital emergency incident responsive mode: Establishment of Novel coronavirus pneumonia department RICU: Respiratory Intensive Care Unit; PCCM: Pulmonary and Critical Care Medicine.

## Results

### 1. High working efficiency

By Mar 02, 2020, we have completed 2974 citywide consultations and treatment of 366 inpatients, including 101 patients diagnosed with COVID-19. Among these patients, the age is 45±18.01, 47 cases are male, and 87 cases are from Hubei province. At the beginning of the NCP department establishment, 5 treatment areas (with 80 doctors and 150 nurses) had been set up in 3 days, and each treatment area can accommodate 17 isolation patients, which ensure all suspected patients in Zhuhai can be isolated and diagnosed in time. Subsequently, total 10 treatment areas (with 162 doctors and 322 nurses) were successively established within one week, which included respiratory intensive care unit (RICU) (for critical patients), 2 severe patients treatment area, 3 common patients treatment area, 2 suspected cases treatment area, one transitional area after nucleic acid removal, and one isolation area before patients discharge.

### 2. Excellent therapeutic effect

Among 101 hospitalized patients with confirmed COVID-19, 23 severe patients had P/F ratio less than 300mmHg, 9 critical patients had P/F ratio less than 150mmHg, accompanied by elevated lactate level. Even if higher rate of severe/critical cases(22.78%), all patients were cured and discharged(Fig 3), except for one death, which had been given invasive mechanical ventilation too early before establishment of NCP department. The knowledge and understood to COVID-19 is constantly supplemented and updated. In NCP department, the professor of respiratory department is directly responsible for the treatment plan, making individualized treatment according to the specific situation of patients, rather than sticking to the guidelines, which is the key to the success of the treatment of rest 100 patients. The effective implementation of the treatment plan benefited from the direct leadership and instruction from the president of hospital. Moreover, due to the strict assurance of discharge evidence, the relapse rate of patients after discharge is only 0.97%. Moreover, no secondary hospital infection, no pipeline infection and no pressure sore were found in severe patients(Fig 3), because 20 doctors and 33 nurses had worked in RICU.

**Figure 3.**
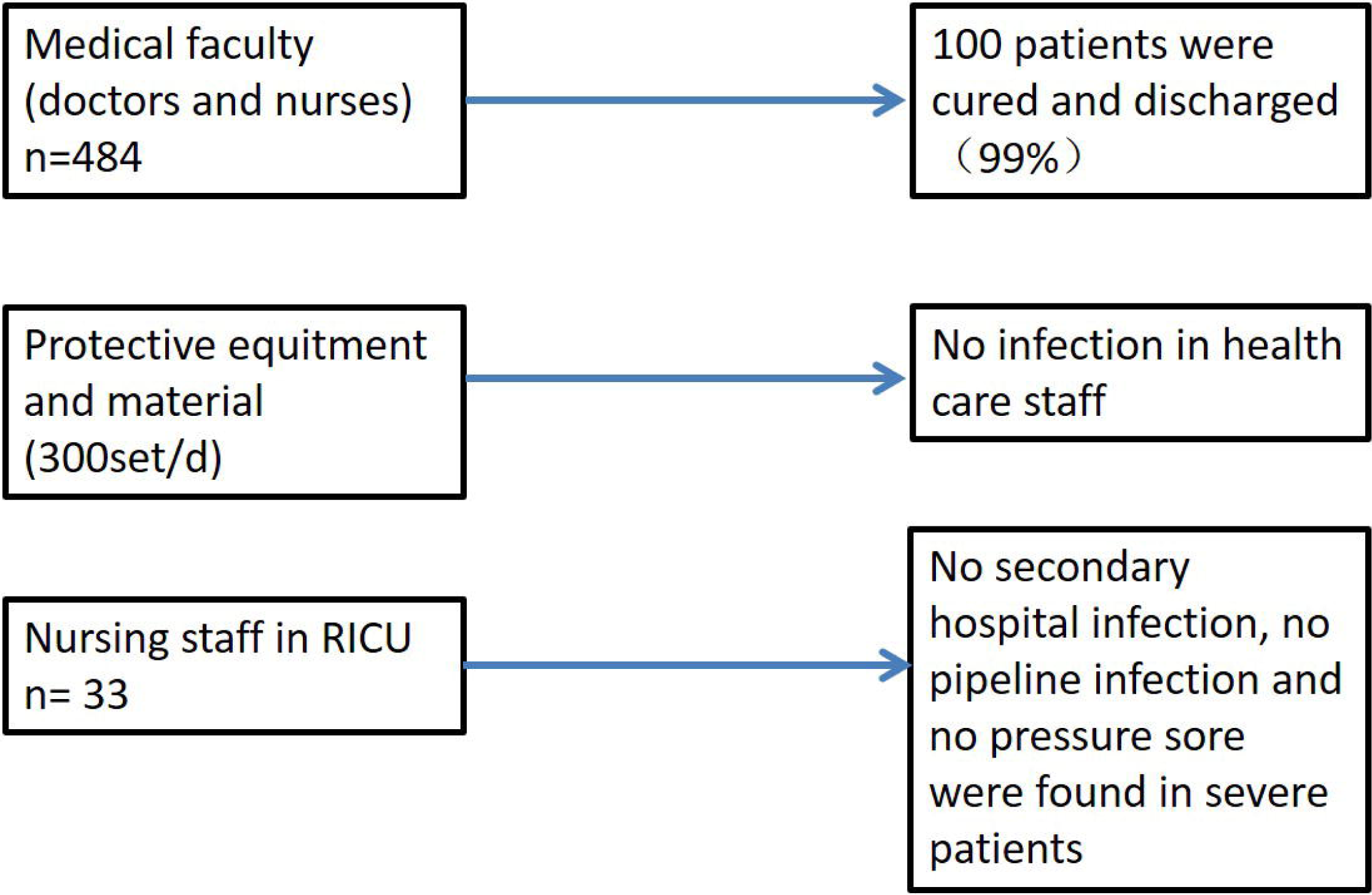
The effect of hospital emergency incident responsive mode in The Fifth Affiliated Hospital of Sun Vat-sen University.

### 3. Finding and confirming person-to-person transmission characteristic of COVID-19 prior to official release conference

Finding, confirming and reporting person-to-person transmission characteristic of COVID-19 prior to official release conference in our hospital. And then the local people’s Health Commission and local administrative departments in Zhuhai were promptly informed, and a timely response plan and stronger protection were set up. The family and close contacts of confirmed COVID-19 patients were isolated quickly and timely, so that the second generation infectious patients (local residents in Zhuhai) was 16, occupying only 15.53% of all patients. Meanwhile, strengthened protection is also key point to zero infection in healthcare group and medical faculty(Fig 3). In NCP department, 300 sets of protective N95 masker and protective suits pro day had been used. And total 1500 protective sets and 9 sets of positive pressure electric air supply respirator had been consumed in RICU.

### 4. Timely adjustment management and treatment strategy prior to guideline update

The first evidence of digestive tract involvement in COVID-19 has been found in our hospital^4^, which improved discharge process of COVID-19 patients. And the earliest clinical trial of chloroquine in the treatment of COVID-19 had been carried out in our hospital, which was helpful for including of chloroquine in updated guideline.

## Discussion

In the face of some public health crisis, there is different emergency system in different country. For example,the Hospital Emergency Incident Command System (HEICS) has emerged as a popular incident command system model for hospital emergency response in the United States and other some countries. Since the inception of the HEICS in 1991, several events have transformed the requirements of hospital emergency management, including the 1995 Tokyo Subway sarin attack, and the 2003 Severe Acute Respiratory Syndrome (SARS) outbreaks in eastern Asia and Toronto, Canada. But HEICS should be continuous improved as additional challenges arise and as hospitals gain further experience with its use^5^. However, in China, due to different political system and other problems, most hospitals still lack the emergency management system similar to HEICS, even if there are some disadvantages in HEICS. In the past 20 years, the major respiratory infectious diseases in China are SARS outbreaks in 2003, H1N1 crisis in 2009, and small-scale prevalent of H7N9 in 2013. In terms of epidemic and transmission scale, this crisis of COVID-19 is close to SARS, so lessons learned from the SARS epidemic should be especially important in responding to the current emergency^6^. In fact, the significant gap in the detection capacity of new infectious diseases and emergency incident responsive system had been found in China^7^. Unfortunately, these gaps are only recognized, but not paid more attention and improved in time, which has become a potential crisis.

There is permanent resident population of about 1.8911 million, but the proportion of floating population is higher and the COVID-19 protection and control task is hard in Zhuhai. Our hospital, the fifth affiliated hospital of Sun Yat-sen University, is the only designated hospitals in Zhuhai. There are advantages and disadvantages to set up designated hospital for management and treatment COVID-19 patients in a comprehensive tertiary hospital. Compared with special infectious disease hospital, comprehensive tertiary hospital has powerful PCCM department, multidisciplinary consultation, and can mobilize more resources. For example, Extracorporeal Membrane Oxygenation (ECMO) was operated by group with the rich experience and thoracic closed drainage and tracheotomy were completed by surgical department successfully in the first patient. On the contrary, too much administrative departments and related clinical departments were involved, resulting in poor implementation in treatment and management, based on traditional hospital responsive mode.

But, our hospital could assess epidemic situation and regulate hospital strategy quickly, establish NCP department with direct leading and management by president of hospital, professional guidance by professor and faculty from PCCM. The infection department was transferred to the periphery in charge of infection control instead of NCP treatment. Depended on rich experience of respiratory specialist on treatment of viral pneumonia, COVID-19 patients had been treated well, particularly for 22 severe and critical patients. Within one month, six versions of the guideline for NCP have been published. Rather than sticking to the guidelines, the opinions from respiratory professor and the expert group had been thorough implemented dependent on individualized treatment, under the direct authorization of the president. Therefore, after the adjustment of the main treatment group and strategy, the overall treatment effect significantly improved, all patients were successfully cured and discharged.

Meanwhile, zero infection in medical faculty was due to early finding of person-to-person transmission characteristic of COVID-19 and enough supply of protective equipment and materials from hospital and government financial support. In addition, in NCP department, under the direct leadership of the president, the scientific research team responded quickly to the problems found in clinical practice. The first evidence of digestive tract involvement in COVID-19 was found by gastroscopy and colonoscopy in our hospital, which improved discharge process of COVID-19 patients. And the earliest application of chloroquine as anti-viral drug dues to direct instruction of president of hospital.

In our hospital, establishment of NCP department, which is administratively led by the president of hospital directly and specialized conduct by respiratory professor, is the key to success in management and treatment of COVID-19 patients. This hospital emergency responsive mode could provide reference for other hospitals and cities in epidemic situation.

## Data Availability

no

## *Ethics approval and consent to participate

Not applicable.

### *Consent for publication

Not applicable.

### *Availability of data and material

All data generated or analysed during this study are included in this published article.

## *Authors’ contributions

Hong Shan and Jing Liu conceived and designed this study and final approval of the version to be published, who were correspondent; Jin Huang and Zhonghe Li performed this study and wrote the manuscript, who contributed equally to this work as first authors; Xiujuan Qu, Xiaobin Zheng and Changli Tu collected and analyzed the data; Meizhu Chen and Cuiyan Tan wrote and embellished this paper. All authors read and approved the final manuscript.

## *Acknowledgements

No

## Reference

1. Situation reports. https://www.who.int/emergencies/diseases/novel-coronavirus-2019/situation-reports.

2. Souza DB, Dall’Agnol CM. Public health emergency: social representations among managers of a university hospital. Rev Lat Am Enfermagem. 2013 Jul-Aug; 21(4): 998–1004. doi: 10.1590/S0104-11692013000400023.

3. Lei Ding, Wei Cai, Jianqing Ding, Xinxin Zhang, Yong Cai, Jianwei Shi, Qiming Liang, Lufa Zhang, Lizhen Sun, Jieming Qu, Fan Jiang, Guoqiang Chen. Ponder over the novel coronavirus infection epidemic situation. SCIENCE CHINA Life Sciences, 2020.2.23 online.

4. Xiao F, Tang M, Zheng X, Li C, He J, Hong Z, Huang S, Zhang Z, Lin X, Fang Z, Lai R, Shan H. Evidence for gastrointestinal infection of SARS-CoV-2. medRxiv. 2020.2.17 online.

5. Arnold JL, Dembry LM, Tsai MC, Dainiak N, Rodoplu U, Schonfeld DJ, Paturas J, Cannon C, Selig S. Recommended Modifications and Applications of the Hospital Emergency Incident Command System for Hospital Emergency Management. Prehosp Disaster Med, 2005 Sep-Oct; 20 (5), 290-300.

6. Cheng VC, Chan JF, To KK, Yuen KY. Clinical management and infection control of SARS: lessons learned. Antiviral Res. 2013 Nov; 100(2):407–19. doi: 10.1016/j.

7. Feng Z, Li W, Varma JK. Gaps remain in China’s ability to detect emerging infectious diseases despite advances since the onset of SARS and avian flu. Health Aff (Millwood). 2011 Jan; 30(1):127–35. doi: 10.1377/hlthaff.2010.0606.

